# Prevalence and Factors Associated with Depression and Anxiety of Hospitalized Patients with COVID-19

**DOI:** 10.1101/2020.03.24.20043075

**Authors:** Xiangyu Kong, Kailian Zheng, Min Tang, Fanyang Kong, Jiahuan Zhou, Le Diao, Shouxin Wu, Piqi Jiao, Tong Su, Yuchao Dong

## Abstract

**Objective:** The 2019 coronavirus disease (COVID-19) epidemic has raised international concern. Mental health is becoming an issue that cannot be ignored in our fight against it. This study aimed to explore the prevalence and factors linked to anxiety and depression in hospitalized patients with COVID-19.

**Methods:** A total of 144 patients diagnosed with COVID-19 were included in this study. We assessed depression and anxiety symptoms using the Hospital Anxiety and Depression Scale (HADS), and social support using the Perceived Social Support Scale (PSSS) among patients at admission. Multivariate linear regression analyses were performed to identify factors associated with symptoms of anxiety and depression.

**Results:** Of the 144 participants, 34.72% and 28.47% patients with COVID-19 had symptoms of anxiety or depression, respectively. The bivariate correlations showed that less social support was correlated with more anxious (r=-0.196, p<0.05) and depressive (r=-0.360,p<0.05) symptoms among patients with COVID-19. The multiple linear regression analysis showed that gender (β=1.446, p=0.034), age (β=0.074, p=0.003), oxygen saturation (β =-2.140, p=0.049), and social support (β =-1.545, p=0.017) were associated with anxiety for COVID-19 patients. Moreover, age (β=0.084, p=0.001), family infection with SARS-CoV-2 (β =1.515, p=0.027) and social support (β =-2.236, p<0.001) were the factors associated with depression.

**Conclusion:** Hospitalized patients with COVID-19 presented features of anxiety and depression. Mental concern and appropriate intervention are essential parts of clinical care for those who are at risk.

## Introduction

Since December, 2019, an outbreak of coronavirus disease 2019 (COVID-19), caused by the severe acute respiratory syndrome coronavirus 2 (SARS-CoV-2) has widely and rapidly spread in China and around the world ^1^. As of March 18, 2020, more than 180,000 confirmed cases and at least 7500 death have been reported in 159 countries (territories/areas), according to the World Health Organization (WHO)^2^. The grim epidemic has caused increasing public panic and mental health stress. Mental health is becoming an issue that cannot be ignored, while trying to control the outbreak.

Previous studies have shown that depression and anxiety are common and persistent mental illness in various chronic diseases^3, 4^, cancer^5^ and other serious illness. And these studies indicated that patients with mental illness, including depression and anxiety may have difficulty with symptom control, and have impaired quality of life. However, recently-published researches on psychological impact of COVID-19 are mainly focus on the healthcare workers^6, 7^ and general public^8^, who were worried about the risks of infection and protective measures, resulting in psychological distress.

Mental health of the hospitalized patients with COVID-19 during the epidemic remains unknown. Considering that patients after diagnosis of COVID-19 were more likely to have psychological concerns such as fear of progression of their illness, disability, or premature death. It is vital to investigate the prevalence and related factors of anxiety and depression in patients infected with COVID-19.

In the present study, we aimed to explore the prevalence and factors linked to anxiety and depression in hospitalized patients with COVID-19. This study may draw more attention to the psychological state of patients with COVID-19, and assist doctors to provide more appropriate treatment and psychological interventions to improve mental and physical health of patients during the campaign to contain and eradicate COVID-19.

## Methods

### Participants

This descriptive quantitative cross-sectional study included 144 patients with confirmed COVID-19 who were admitted from 23 February 2020 to 13 March 2020 in Huoshenshan Hospital (Wuhan, China) during the COVID-19 epidemic. All patients were diagnosed with COVID-19 according to World Health Organization interim guidance. Informed consent was provided by subjects before study commencement. All participants completed the questionnaires through an online survey platform (‘SurveyStar’, Changsha Ranxing Science and Technology, Shanghai, China) at admission. The study was approved by the Research Ethics Commission of Huoshenshan Hospital.

### Hospital Anxiety and Depression Scale (HADS)

The HADS is a self-assessment questionnaire designed to detect anxiety and depression symptoms in general hospital patients. It has been acknowledged that the Chinese version of the HADS had good internal consistency and favorable scale equivalence^9^. The degree of anxiety and depression is rated by the accumulated scores: score 0-7, indicating no anxiety or depression; score 8-10, indicating mild levels of anxiety or depression; score 11-14, indicating moderate and score 15-21, indicating severe levels of anxiety or depression.

### Perceived Social Support Scale

The 12-item Perceived Social Support Scale (PSSS) were translated into Chinese and has been widely adopted to measures perceived support from family, friends and other ways in Chinese population^10^. Total scores range from 0 to 84, with higher scores implying greater level of perceived social support.

### Statistical Analysis

SPSS software, version 19 were used for statistical analysis. Means and proportions of the given data for each variable were calculated. Categorical variables were analyzed using the Pearson’s chi-square test or Fisher’s exact test. Continuous variables were analyzed using non-paired Student *t* test or one-way analysis of variance (ANOVA). Multivariate regression analysis was performed to identify factors associated with depression and anxiety. Differences between groups were considered to be significant when the P-value was < 0.05.

## Results

### Demographic characteristics

A total of 144 participants, including 70 male and 74 female were included in current study. The age of the study participants ranged from 15 to 87 years. Their average age was 49.98±13.73 years. Participants were mostly living with a spouse (121/144, 84%). About a third of the subjects (54/144, 37.5%) were well educated (≥ bachelor’s degree), and only 4 of 144 participants (2.8%) had primary education. Oxygen saturation is a key clinical index for evaluation the severity of patients with COVID-19. In the present study, 11.1% participants whose oxygen saturation was less than or equal to 93% at rest, were with severe disease. Considering that other family members’ infection may cause emotional distress to the participants, we also collected the infection status of family members. 59 participants (41%) had one or more family members infected. Demographic characteristics are listed in Table 1.

**Table 1.** Baseline demographic and clinical characteristic of patients with COVID-19.

|  | n | % |
| --- | --- | --- |
| Gender |  |  |
| Male | 70 | 48.6 |
| Female | 74 | 51.4 |
| Age (years) |  |  |
| ≤50 | 70 | 48.6 |
| >50 | 74 | 51.4 |
| Marital Status |  |  |
| Married | 121 | 84.0 |
| Single | 17 | 11.8 |
| Divorced | 2 | 1.4 |
| Widowed | 4 | 2.8 |
| Education Status |  |  |
| Primary | 4 | 2.8 |
| Lower secondary | 34 | 23.6 |
| Upper secondary | 52 | 36.1 |
| University/master/doctorate | 54 | 37.5 |
| Oxygen saturation at rest |  |  |
| ≤93% | 16 | 11.1 |
| >93% | 128 | 88.9 |
| Infection status of family members |  |  |
| Infected | 59 | 41.0 |
| Non-infected | 85 | 59.0 |

### Psychosocial characteristics of the participants with COVID-19

The mean score of anxiety subscale and depression subscale for all patients was 6.35±4.29 and 5.44 ± 4.32, respectively. With the reference to HADS, 50 (34.72%) and 31 (28.47%) participants presented symptoms of anxiety and depression, respectively. Regarding the patients’ anxiety levels, it was found that 17.36%, 12.5% and 4.86% appeared to have mild, moderate and severe anxiety, respectively. As for the depression levels of patients, 20 were mildly depressed (13.89%), 15 were moderately depressed (10.42%), and 6 were severely depressed (4.17%).

### Correlations among depression, anxiety and social support in COVID-19 patients

Since there is a large body of evidence that social support plays a beneficial role in mental health^11^. Self-reported levels of social support were assessed among the patients with COVID-19. The mean social support score for all participants was 63.41±11.99. The average score of family, friends and other support was 22.35±4.42, 20.53±4.60 and 20.52±4.55, respectively. More than half of the participants (90/144, 62.5%) exhibited high level of perceived social support.

The bivariate correlations showed that less social support was correlated with more anxious (r=-0.196, p<0.05) and depressive (r=-0.360,p<0.05) symptoms (Table 2). In detail, friend support (r = -0.165, p < 0.05) and other support (r=-0.230, p < 0.05) were significantly negative correlated with anxiety. In addition, family support (r=-0.283, p < 0.05), friend support (r=-0.307, p < 0.05) and other support (r=-0.363, p < 0.05) were significantly negative correlated with depression.

**Table 2.**
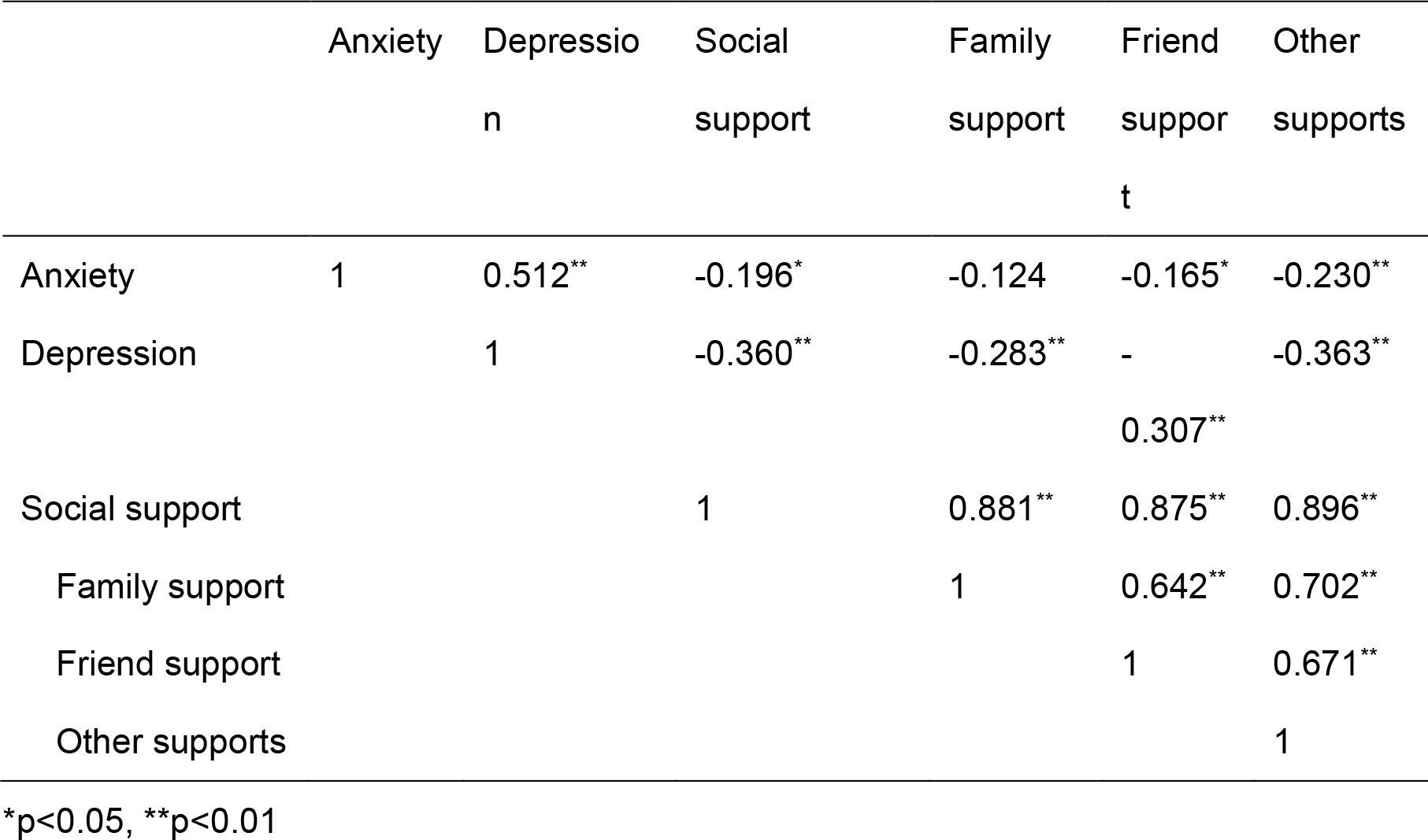
Association between anxiety, depression and social support.

### Factors associated with depression and anxiety among patients with COVID-19

In order to investigate the factors related to depression and anxiety among patients with COVID-19, anxiety and depression scores were compared between different groups. As shown in Table 3, anxiety and depression scores were significantly higher in those who are older (age>50) and low-educated. Additionally, patients with lower oxygen saturation had higher anxiety score, and those getting less social support had higher depression scores.

**Table 3.** Comparison of anxiety and depression scores on different variables.

|  | Anxiety score<br>(mean±SD) | p | Depression score<br>(mean±SD) | p |
| --- | --- | --- | --- | --- |
| Gender |  |  |  |  |
| Male | 5.71±3.98 | 0.082 | 5.47±4.30 | 0.942 |
| Female | 6.96±4.51 |  | 5.42±4.39 |  |
| Age (years) |  |  |  |  |
| ≤50 | 4.91±3.40 | **<0.001** | 4.33±4.44 | **0.002** |
| >50 | 7.72±4.62 |  | 6.50±3.97 |  |
| Marital status |  |  |  |  |
| Married | 6.53±4.24 | 0.264 | 5.50±4.13 | 0.745 |
| Single/divorced/<br>widowed | 5.43±4.54 |  | 5.17±5.36 |  |
| Education level |  |  |  |  |
| Primary/secondary | 7.09±4.66 | **0.008** | 6.06±4.47 | **0.028** |
| University/master/<br>doctorate | 5.13±3.30 |  | 4.43±3.92 |  |
| Oxygen saturation at rest |  |  |  |  |
| ≤93% | 8.75±5.88 | **0.017** | 6.50±5.53 | 0.303 |
| >93% | 6.05±3.98 |  | 5.31±4.16 |  |
| Infection status of<br>family members |  |  |  |  |
| Infected | 6.92±4.33 | 0.192 | 6.19±4.63 | 0.087 |
| Non-infected | 5.96±4.25 |  | 4.93±4.05 |  |
| Social support |  |  |  |  |
| High | 5.89±4.28 | 0.093 | 4.53±3.89 | **0.001** |
| Low-Moderate | 7.13±4.23 |  | 6.96±4.63 |  |

The multiple linear regression analysis (Table 4) showed that gender (β=1.446, p=0.034), age (β=0.074, p=0.003), oxygen saturation (β =-2.140, p=0.049), and social support (β =-1.545, p=0.017) were associated with anxiety for COVID-19 patients. It suggested that female, and patients who are older, with lower oxygen saturation and less social support would tend to present anxiety symptoms. Moreover, age (β=0.084, p=0.001), family infection with SARS-CoV-2 (β =1.515, p=0.027) and social support (β =-2.236, p<0.001) were the factors associated with depression. The results indicated that patients with older age, family member infection and less social support were more likely to be depressive (table 4).

**Table 4.**
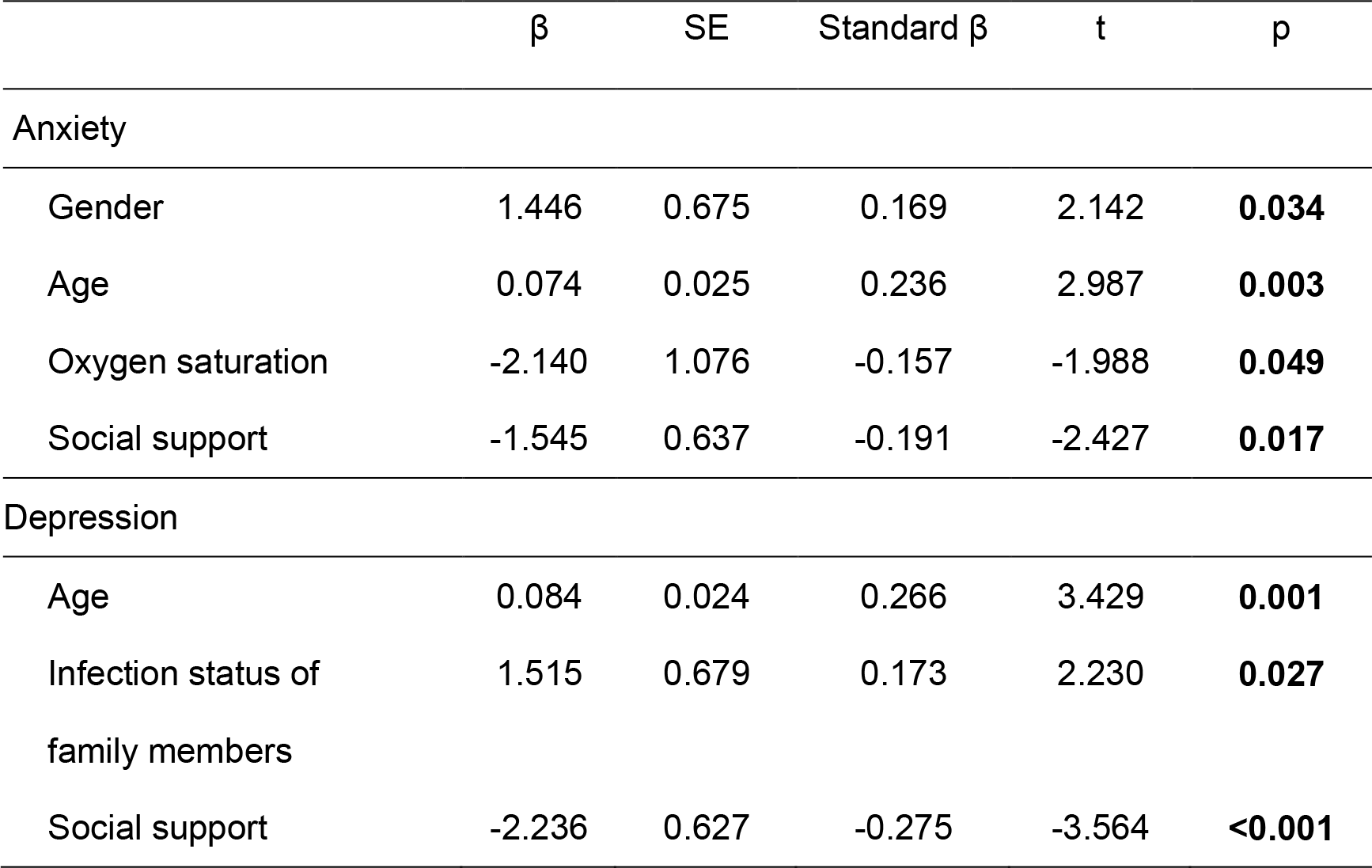
Multivariate regression analysis of factors associated with anxiety and depression.

| | $\beta$ | SE | Standard $\beta$ | t | p |
| --- | --- | --- | --- | --- | --- |
| Anxiety |  |  |  |  |  |
| Gender | 1.446 | 0.675 | 0.169 | 2.142 | **0.034** |
| Age | 0.074 | 0.025 | 0.236 | 2.987 | **0.003** |
| Oxygen saturation | -2.140 | 1.076 | -0.157 | -1.988 | **0.049** |
| Social support | -1.545 | 0.637 | -0.191 | -2.427 | **0.017** |
| Depression |  |  |  |  |  |
| Age | 0.084 | 0.024 | 0.266 | 3.429 | **0.001** |
| Infection status of<br>family members | 1.515 | 0.679 | 0.173 | 2.230 | **0.027** |
| Social support | -2.236 | 0.627 | -0.275 | -3.564 | **<0.001** |

## Discussion

A number of studies have interlinked the depression and anxiety to patients with different disease^3-5^. This study first reported the prevalence of anxiety and depression in patients with COVID-19 during the epidemic. The results of the present study showed that 34.72% and 28.47% patients with COVID-19 had symptoms of anxiety or depression, respectively.

In the present study, it is worthy of note that social support is one of the key factors linked to anxiety and depression for patients with COVID-19 (Table 4). As the results displayed that less social support was correlated with more anxious and depressive symptoms (Table 2). Mounts of studies have demonstrated that in the case of disease, patients need more social support, which included physical and psychological assistance provided by family members, friends, medical workers, and relevant institutions to cope with difficulty^11^. There is consistent evidence which showed that social isolation and loneliness were linked to worse mental health outcomes^12^. While during the COVID-19 epidemic, many isolated patients often felt helpless and lonely due to the lack of family or friends accompany. In the circumstances, medical workers as the major peer support that is of great significance to infected patients. In clinical practice, Chinese medical members would keep in touch with patients and try various psychological support methods to help isolated patients rebuild confidence. In some Wuhan makeshift hospitals, patients with mild symptoms did Tai Chi practice (which has been verified as an effective way to improve lung function for COPD patients^13^), singing and dancing as physical relax, accompanied and guided by medical staff. This kind of doctor-patient interaction may encourage patients to stay positive mindset.

Furthermore, older age and lower oxygen saturation are the other factors should be considered for patients to be anxious. As previous research revealed that older patients are at increased risk with severe COVID-19 symptoms and death ^14^. Additionally, oxygen saturation is a key index to evaluate the severity of patients with COVID-19. According to the Chinese management guideline for COVID-19 (version 6.0)^15^, patients whose oxygen saturation was≤93% at rest was defined as severe type patients. In this study, 11.1% participants were with low oxygen saturation. These results indicated that patients with severe illness are more likely to be anxious. More psychological care and health attention needs to be given to these critically ill patients.

Consistent with previous report, which is focus on the psychological responses among general population during the COVID-19 epidemic in China^8^, females patients are also prone to developing higher levels of anxiety as shown in the current study. Meanwhile, education background is another associated factor to the mental distress among infected patients. And as we expected, family member infection is another factor affecting patients to be depressed. High levels of concern about other family members and lack of family care may magnify pessimism over the illness.

Given that anxiety and depression are related with longer hospitalization^3, 16^ and non-adherence to treatment^17, 18^ in several diseases. Early prevention of mental health problems is of vital importance to help patients have good clinical outcomes and better life quality. As the COVID-19 epidemic continues to spread, our findings is particularly instructive to develop a psychological support strategy for hospitalized patients with COVID-19 in China and other areas affected by the epidemic.

### Study Limitation

1) The present study was single-centered, the study sample was not representative of all patients with COVID-19 in China, which limits the generalizability of the results. 2) We only collected data at admission. Thus, it did not allow for investigation the inferences or changes over time. 3) This cross-sectional study is not capable to determine a causal relationship between mental health (anxiety or depression) and the sociodemographic and clinical variables.

## Conclusion

Hospitalized patients with COVID-19 experienced features of anxiety and depression. The significant factors found in the present study may draw medical workers paying more attention to the mental health of patients with COVID-19, and psychological care and timely intervention are needed for patients during the epidemic.

## Data Availability

The data used to support the findings of this study are included within the article.

